# Remdesivir to treat COVID-19: can dosing be optimized?

**DOI:** 10.1101/2021.06.16.21258981

**Authors:** Jessica M. Conway, Pia Abel zur Wiesch

## Abstract

The antiviral remdesivir has been approved by regulatory bodies such as EMA and FDA for the treatment of COVID-19. However, its efficacy is debated and toxicity concerns might limit the therapeutic range of this drug. Computational models that aid in balancing efficacy and toxicity would be of great help. Parametrizing models is difficult because the prodrug remdesivir is metabolized to its active form (RDV-TP) upon cell entry, which complicates dose-activity relationships.

Here, we employ a computational model that allows predicting drug efficacy based on the binding affinity of RDV-TP for its target polymerase in SARS-CoV-2. We identify an optimal infusion rate to maximize remdesivir efficacy. We also assess drug efficacy in suppressing both wild-type and resistant strains, and thereby selection of resistance.

Our results differ from predictions using prodrug dose-response curves (pseudo-EC_50_s). We expect that reaching 90% inhibition (EC_90_) is insufficient to suppress SARS-CoV-2 in lungs. While standard dosing mildly inhibits viral polymerase and therefore likely reduces morbidity, we also expect selection for resistant mutants for most realistic parameter ranges. To increase efficacy and safeguard against resistance, we recommend continuing remdesivir use with companion antivirals and/or with dosing regimens that substantially increase the levels of RDV-TP.

## 1. Introduction

In November 2020, the first ACTT-1 study results were released showing that the antiviral drug remdesivir (commercial name Veklury) had some efficacy in treating COVID-19, specifically in shortening the time to recovery in adults who were hospitalized with COVID-19 and had evidence of lower respiratory tract infection [1]. It was mid-pandemic and as there were, at the time, no therapeutic agents shown to be efficacious against COVID-19, study results were very welcome news. Remdesivir had already been granted emergency usage approval by large organizations such as the European Medical Association (EMA)[2] and the US Food and Drug Administration (FDA)[3]. This approval was granted based largely on evidence that remdesivir successfully prevented disease in rhesus macaques infected with Middle East respiratory syndrome coronavirus (MERS-CoV), which is closely related to SARS-CoV-2, the virus associated with COVID-19 [4] and preliminary results ACTT study results showing remdesivir accelerated recovery from advanced COVID-19 [5].

In spite of these positive results, the efficacy of remdesivir remained a topic of debate. Significantly, a follow-up WHO meta-analysis – for the Solidarity Clinical trial - suggested that remdesivir is not effective [6]. However, questions surrounding study selection and mathematical approach in this analysis remain, and a more focused meta-analysis of the same data shows some benefit in patients requiring oxygen [7]. More recent meta-analyses and studies show that remdesivir improves recovery time [7,8] but does not reduce mortality [7]. These are observation studies, but one can also, with some difficulty, investigate the impact of remdesivir on SARS-CoV-2 viral replication in host. While a modeling study from the French COVID Cohort Investigators, and French Cohort Study groups suggested remdesivir did not improve SARS-CoV-2 viral clearance time [9], the sample size was small, animal [10] and larger human studies [11] do show accelerated viral decay in remdesivir-treated macaques and humans, respectively.

Remdesivir efficacy remains unclear, and questions of whether it should be abandoned, or optimized for use in combination therapy for COVID-19, remains. On the strength of the encouraging, though not definitive, monotherapy studies, follow-up studies are investigating remdesivir in combination with other therapies [12], with positive results. For example results from the ACTT-2 study showed that remdesivir in combination with baricitinib, a Janus kinase inhibitor offered significant improvements in reducing recovery time and accelerating clinical improvement [13]. Multiple studies investigating remdesivir in combination with other drugs, such as dexamethasone [14], are ongoing.

The major challenge is that remdesivir dosing itself has not been optimized. Study outcomes showing poor efficacy may be a consequence of sub-optimal dosing. At the time of this writing, we are not aware of dose-ranging or dose fractionation studies for more effective RDV regimens. Here we propose to predict optimal RDV regimen through pharmacokinetic and pharmacodynamic (PK/PD) modeling. PK/PD modeling is a cornerstone of optimizing drug dosing. The metabolic pathway leading from extracellular RDV to intracellular RDV-TP is not yet confirmed [15–17]. However, there is consensus that intracellular RDV-TP is the active form of the drug [16–18]. For the purpose of this study, we therefore a novel PK model to describe the distribution in the body of the remdesivir (RDV) prodrug and its active triphosphate metabolite GS-443902, (RDV-TP), parametrized using data from safety and tolerability investigations reported in Humeniuk et al. (2020) [15]. Our PK model distinguishes itself from the work by Goyal et al. [10] in its emphasis on RDV-TP, which has a longer half-life than RDV and therefore alters dosing recommendations significantly.

We further develop a novel PD model that relies on the underlying interactions between RDV-TP and target receptor to predict a drug regimen’s efficacy as the drug interferes with viral replication. In general, incorporating drug-target binding in models has added to our understanding of drug therapy and was shown to improve quantitative predictions of drug action in antibiotic and HIV therapy [19–23]. Optimizing dosing therefore increasingly relies not on PK/PD models, but PK/TE/PD models where TE stands for target engagement [24].

The PK/TE/PD models offer a complementary approach to models employing allometry and mouse models. It is common practice to extrapolate from mouse models to humans, and this approach is very helpful if the target tissue is not easily accessible in human patients (e.g. the lungs) but can be obtained from animal models. Indeed this approach was taken in Hanafin et al. [25], aiming to identify improved RDV dosing strategies. Their allometry-based modeling predicted that intravenously-delivered RDV could not achieve 90% maximal inhibitory concentration, as measured *in vitro*, of unbound remdesivir in human plasma. While the strategy is common and sound, drugs that that undergo extensive metabolism, which is the case for RDV [15,16], can override the effect of size in simple scaling of drug doses [26]. In addition, reliable allometric scaling to predict human doses typically involves more than one model species [27]. We conclude that our PK/PD model, relying on pharmacokinetic measurements of RDV and RDV-TP, should deliver more accurate predictions.

We use our model to investigate efficacy of current drug regimen and to predict drug regimen with improved efficacy, while being mindful of the possibility of drug toxicity. We then investigate the propensity of proposed drug regimen to engender and select for drug resistance in SARS-CoV-2. Drug resistance is a major obstacle to delivering effective antiviral or antibiotic therapy for viral or bacterial infections respectively. The driving motivation behind the use of a combination of antiretroviral drugs to treat HIV, for example, is prevention of drug resistance [28]. In the worst case we lose effective therapies. For example, amantadanes (amantadine and rimantadine) were very effective in treating influenza A, with efficacy rates of up to 90%. While in use from the 1960s to the 2000s, there was a significant uptick in resistant circulating influenza strains in the 2000s, with more than 69% of H1 subtypes resistant as of 2020. It is precisely due to these high level of resistance that adamantanes are no longer recommended for treatment of influenza A [29].

Already there is documentation of emergent RDV resistant SARS-CoV-2 in an immunocompromised patient being treated with RDV for COVID-19 [30]. This observation matches expectations from *in vitro* studies [31,32]. Specifically, RDV inhibits the SARS-CoV-2 RNA-dependent RNA polymerase (RdRp, encoded by *nsp12-nsp7-nsp8*), and mutations in RdRp can decrease sensitivity to RDV. What is most troubling is that, *in vitro*, these mutations leave viral fitness largely unaffected [31–33]. Thus as we will show, resistance can easily be selected for by some RDV drug regimen. While at this time there is limited concern that resistant strains will transmit or become dominant, as is the case for amantadane-resistant strains of influenza A, since RDV is administered in hospital only, acquired RDV resistance of SARS-CoV-2 in a host can limit the capacity of RDV to accelerate viral clearance and improve clinical outcomes.

In the following, we describe our model of PK and PD of RDV and its intracellular active metabolite RDV-TP. Referencing estimates for rates of viral replication in the lower respiratory tract (LRT) and upper respiratory tract (URT) [34], we show efficacy of current treatment regimen. Note that since we parametrize our model using PK parameters from plasma rather than tissues [25], our results represent a best-case-scenario. We then derive from our model an optimal dosing rate and show that at this rate, one can achieve RDV-TP concentrations sufficient to fully suppress spread within the LRT, but not the URT. We therefore conclude that RDV will serve best as a companion therapy. Finally we show the importance of careful dosing by demonstrating our model predictions on the broad regime of drug regimen that will select for RDV resistant variants.

## 2. Materials and Methods

### 2.1 Pharmacokinetic model

We develop a PK model to describe the distribution in the body of the remdesivir (RDV) prodrug and its active triphosphate metabolite GS-443902, (RDV-TP) parametrized using data from Humeniuk et al. (2020) [15]. We focus on these two components because the metabolic pathway leading from extracellular RDV to intracellular RDV-TP is not yet confirmed [15–17]. However, there is consensus that intracellular RDV-TP is the active form of the drug [16–18].

Examination of the available data suggests nonlinear behavior. Notably, RDV shows biphasic decay, with a rapid drop in concentration following termination of RDV infusion. Further, reportedly the half-life of RDV-TP in PBMCs varies with dose size and duration of infusion, as does the peak concentration (note that, to our knowledge, there is no available longitudinal data on GS-443902). We are therefore motivated to develop a nonlinear model to describe their dynamics. The model equations are given in equation (1), where *R* represents the concentration of remdesivir (RDV) in plasma, *P* in the periphery, and *A* represents the intracellular concentration of the active metabolite RDV-TP.

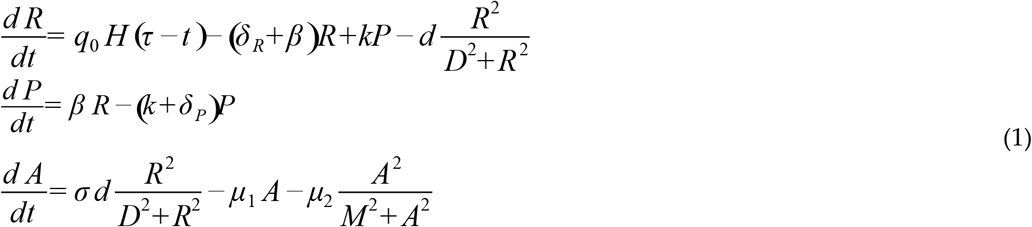

Remdesivir is infused at a constant rate with duration ^*τ*^, *q*_0_= *Dose* / *τ* where *τ* is the infusion duration. We model the short infusion using a Heaviside function,

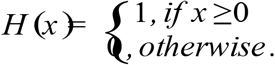

RDV in plasma, *R*, diffuses to the periphery at rate *β*, returns from the periphery at rate *k*, and is eliminated at rate *δ R*. RDV in the periphery, *P*, returns is eliminated at rate. RDV in plasma, *R*, is also absorbed into cells and converted through a series of reactions to the active metabolite *A*. In absence of definitive reactions and longitudinal data on intermittent steps, we model this process using a Hill function with rate *d* and Hill coefficient 2. *A* is eliminated linearly at rate *µ*_1_ and nonlinearly at rate, again modeled with a Hill function with Hill coefficient 2.

We estimate model parameters using the Nelder-Mead algorithm as implemented by the “optim” function in R. Specifically we use longitudinal RDV concentration measurements in the plasma following single-dose administration ([15], Figure 1 and Figure 2) in addition to PBMC pharmacokinetic parameters (*AUC*_*∞*_, C_max_, C_24_, and t_1/2_) of RDV-TP in the single-ascending-dose study in Humeniuk et al. [15]. Parameter estimates are provided in Table 1 and details on the fitting are provided in Supplementary Materials Text S1. We show how our model fits compare with the data in Figure 1 and Supplementary Materials Figure S1 and Table S1.

**Figure 1.**
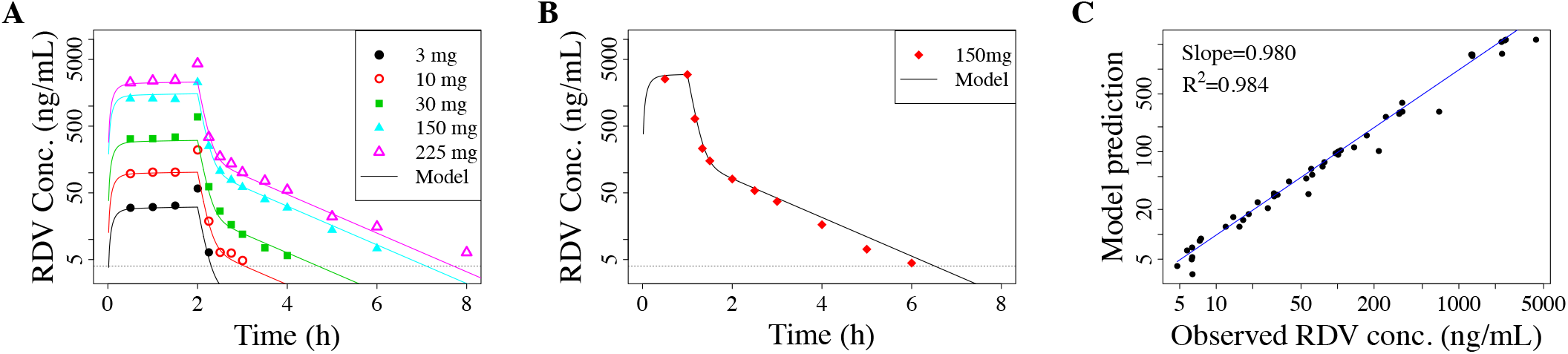
Median RDV plasma concentration from single-dose experiments [15], with different RDV dose size and infusion duration, compared to our pharmacokinetic model prediction (equation (1), main text) given (**a**) 2-hour infusions of 3 mg, 10 mg, 30 mg, 75 mg, 150 mg, 225 mg and (**b**) 1-hour infusion of 150 mg. (**c**) Linear regression comparing the model predictions with the data to demonstrate how well the model explains the data. Data from 75 mg dosing excluded in the fitting, see Supplementary Materials Text S1 for details and Figure S1 for comparison. For cases where Humeniuk et al. (2020) [15] gives plasma concentrations following multi-dose administration, we use the first dose only.

**Figure 2.**
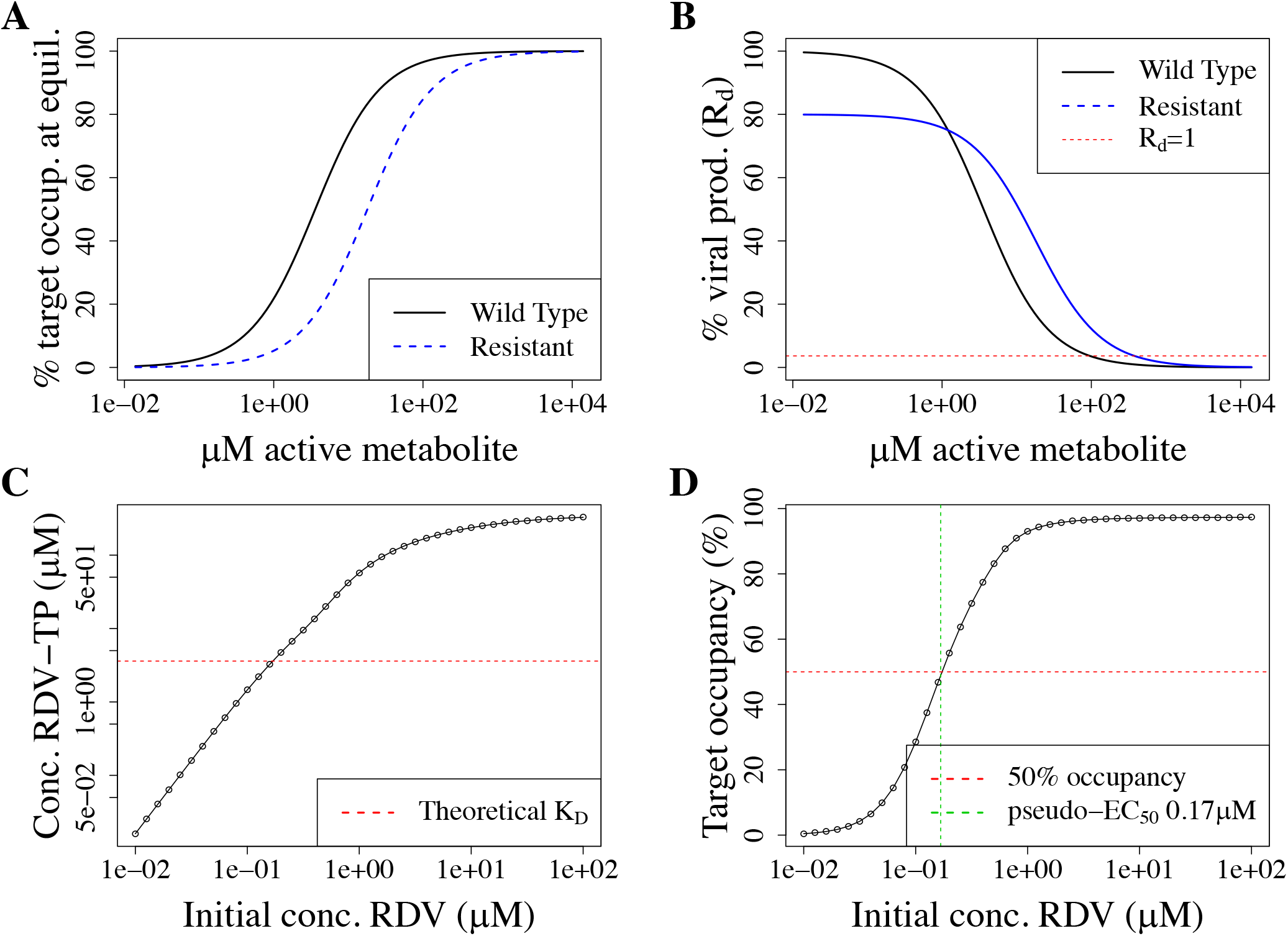
Pharmacodynamic model. (**a**) Target occupancy at equilibrium depending on RDV-TP concentration as calculated with equation (2). We assume that a resistant strain has a five-fold reduced K_D_. (**b**) Resulting viral production rate (and therefore R_d_, see equation (3)) relative to the rate in absence of drugs. Here, we assume the same parameters as in (A), and additionally set a fitness cost for the resistant strain of 20%. (**c**) & (**d**): Model validation by simulating “pseudo-EC_50_” for remdesivir in vitro. This figure shows simulations of in vitro experiments to determine the half-maximal suppression of viral production (EC_50_) in cell culture when supplying RDV to the medium of cells grown in culture (i.e. extracellular RDV). (**a**) Intracellular RDV-TP concentrations depending on the extracellular RDV concentration (black line) obtained with our pharmacokinetic model described in section 2.1. The dashed red line indicates the K_D_ of RDV-TP to polymerase binding. (**b**) Target occupancy depending on extracellular RDV concentrations obtained with equation (2) (black line). The dashed red line indicates 50% target occupancy. The dashed green line indicates the EC_50_ as obtained by fitting a log-logistic function to the calculated data shown in this figure with the R package “drc”.

**Table 1:** Pharmacokinetic model parameter estimates. The parameters β and k also implicitly include the transition with apparent volumes since the ODEs represent dynamics of drug concentrations. That is, *k*=(drug transport rate)* V_p_/V_r_.

| Parameter | Description | Units | Value |
| --- | --- | --- | --- |
| *Remdesivir* |  |  |  |
| Dose | Total mass of RDV infused | mg | varies |
| $\tau$ | Duration of infusion | h | varies |
| $V_r$ | Apparent volume | L | 6.09 |
| $\delta_R$ | Elimination rate | per hour | 4.16 |
| $k$ | Periphery-to-plasma RDV concentration transition rate<sup>1</sup> | per hour | 0.13 |
| $\beta$ | Plasma-to-periphery RDV concentration transition rate | per hour | 4.42 |
| $d$ | Nonlinear plasma RDV to RDV-TP (intracellular GS-443902) transport and activation rate | mg/L/h | 31.57 |
| $\sigma$ | Conversion of concentrations from mg/L to $\mu\text{M}$ | mol/mg | 1 |
| $D$ | RDV concentration yielding 50% of the maximal transport/activation rate | mg/L | 3516.27 |
| *Remdesivir concentration in the periphery* |  |  |  |
| $\delta_P$ | Elimination rate | per hour | 0.61 |
| *Intracellular GS-433902 (RDV-TP)* |  |  |  |
| $\mu_1$ | Linear elimination rate of GS-433902 | per hour | 1.55 |
| $\mu_2$ | Nonlinear elimination rate of GS-433902 | $\mu\text{M}/\text{h}$ | $8.38 \times 10^{-3}$ |
| $M$ | RDV-TP concentration at which the nonlinear elimination rate is 50% its maximum | $\mu\text{M}$ | 17.19 |

Note that while we estimate model parameters from single-dose studies, we model multiple-dose drug regimen. We justify this use by noting that there was no observed accumulation of RDV in the multi-dose study, likely because of its short half-life [15]. There is no data on the metabolite RDV-TP from multi-dose studies, but we note that Humeniuk et al. 2020 [15] reported accumulation of an intermediate metabolite, and our nonlinear model predictions also show such an accumulation.

### 2.2 Pharmacodynamic model

To the best of our knowledge, the IC_50_ or EC_50_ of RDV-TP has never been determined experimentally. We therefore assume that the production of new virions is inversely proportional to the number of occupied binding sites in the viral target, the polymerases producing a nascent RNA chain. The half-life of intracellular RDV-TP is very long, such that the binding should be in equilibrium and the equilibrium binding constant K_D_ is sufficient to calculate target occupancy. In the absence of other data, we use the predicted K_D_ as derived via the binding energy from molecular modeling studies (K_D_=3.6μM, [35]). In HCV, the directly measured IC_50_ of RDV-TP was close to this value (5.6 µM) [36]. *Importantly, we here define the target as all potential insertion sites where RDV-TP could integrate into the nascent RNA chain and thereby disrupts the production of a functional viral genome. In reality this process is very complicated* [36]. *Here, we use a very simplified approach to be able to work with the available data-for more complicated approaches, we do not have enough reliable parameter measurements yet*. Thus, we assume that the antiviral efficacy of RDV-TP is at its EC_50_ when the intracellular concentration is exactly at the K_D_.

The fraction of occupied target, f_occupied_, is then described by

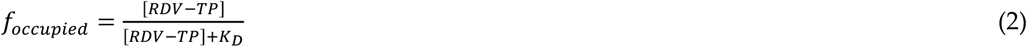

where [RDV-TP] is the concentration of RDV-TP and K_D_ the affinity as described above. As stated above, we assume here that the production rate of new virions, is inversely proportional to f_occupied._ In standard models of viral within-host dynamics, R_0_, the number of new cells an infected cell infects, is proportional to the viral production rate [37]. We therefore assume that the effective viral reproductive number depending on the drug concentration, R_d_, depends on R_0_ and the amount of free target:

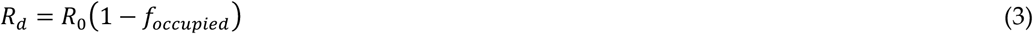

To suppress viral spread between cells, R_0_ has to be below 1. Thus, the minimal effective fraction of occupied target becomes

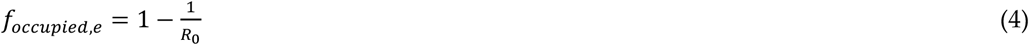

### 2.3 Concentrations selecting for resistance

Resistant strains have a lower effective affinity, either because mutations lead to reduced binding, or because excision repair removes RDV-TP from the nascent RNA chain. Since the affinity constant K_D_ is inversely correlated to affinity, the effective K_D_,_res_ of the resistant strain must be larger than the one for the wild-type, K_D_,_wt_. This results in a lower target occupancy for the resistant than the wt strain at the same concentrations of RDV-TP (Figure 2a).

At the same time, resistant mutants are typically less fit than the wild type and their within-host R_0_ is lowered by w_res_, which describes the relative fitness of the resistant mutant and *R*_0,*res*_= *w*_*res*_ *R*_0,*wt*_.

Using equation (2) and (3) to obtain the minimal effective concentration for both wt and resistant strain, we then obtain

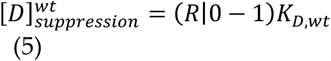

This means that less drug is needed when K_D_ becomes small (and therefore drug-target affinity large), and more drug is needed when R_0_ rises (and therefore the virus replicates more).

The minimal effective concentration for resistant strains is given by

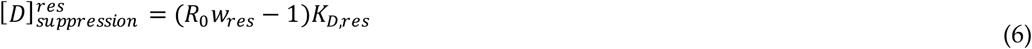

The latter is also the highest dose that still selects for resistance, i.e. the upper limit of the resistance-selection window in Figure 2b.

The lower limit of the resistance selection window is given by the lowest concentration at which the viral production rate and therefore R_D_ of the resistant strain exceeds the one of the wild-type. With *K*_*D,res*_ = *K*_*D*,_*k*_*res*_ where k_res_ describes the decrease in effective drug-target affinity in the resistant strain, this concentration, [D]_selection_, is given by

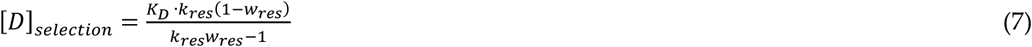

To be selected for at a given drug concentration 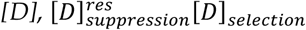 i.e. there must be at least one concentration at which the resistant strain is selected for but not suppressed yet. For a given decrease in effective drug-target affinity, the minimal fitness of a resistant strain that allows resistance to arise can therefore be given by

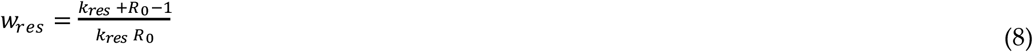

### 2.4 Model validation: recapitulation of in vitro experiments

While the dose-response curve of RDV-TP has never been measured in vitro, there are several publications that investigated the dose-response relationship and EC_50_ of the parent drug remdesivir in vitro as measured by the inhibition of viral production. This has also been referred to as a “pseudo-EC_50_” because it does not reflect the molecular mechanism of action. Additionally, remdesivir is thermally unstable and rapidly degraded at 37 °C [38], such that the initial remdesivir concentration is not suitable for obtaining an EC_50_. As a validation step, we set out to investigate whether our PK model (that describes the metabolization of RDV to RDV-TP and cell entry) together with our PD model can reproduce realistic estimates for the pseudo-EC_50_ (Figure 2 and Supplementary Materials Figure S2). It is important to note that our PK model was fitted to RDV-TP levels in PBMCs, which are not primary targets for SARS-CoV-2 but a proxy for intracellular concentrations because they can be easily obtained. Also, EC_50_s and RDV-TP content strongly depend on the cell type [39]. However, our estimate of 0.17 µM is well within the range of reported pseudo-EC_50_ values of 0.01 µM in isolated human epithelial airway cells, 0.28 µM in Calu cells and 1.65 µM or 0.77 µM in Vero E6 cells [39,40].

## 3. Results

### 3.1 Standard dosing does not achieve effective concentrations

SARS-CoV-2 can replicate in many different body compartments, and its ability to replicate likely differs between those compartments. In non-systemic infections, the replication rate in the upper and lower respiratory tract (URT and LRT) are most important for understanding disease outcome. R_0_ has been estimated to be 8.5 [4.3-25] for the URT and 27.5 [19.8-42.1] for the LRT [34]. Using equations (2) and (3), we therefore assume that a target occupancy of on average 88% [77-96]% in the URT and 96% [95-98]% in the LRT is required to suppress viral replication. Figure 3a illustrates the pharmacokinetics of RDV and RDV-TP under standard RDV dosing (30 min-2 h infusion of a loading dose of 200 mg and 100 mg thereafter [1]. Figure 3b illustrates the resulting target occupancy and indicates the minimal effective target occupancy as described above for both URT and LRT.

**Figure 3.**
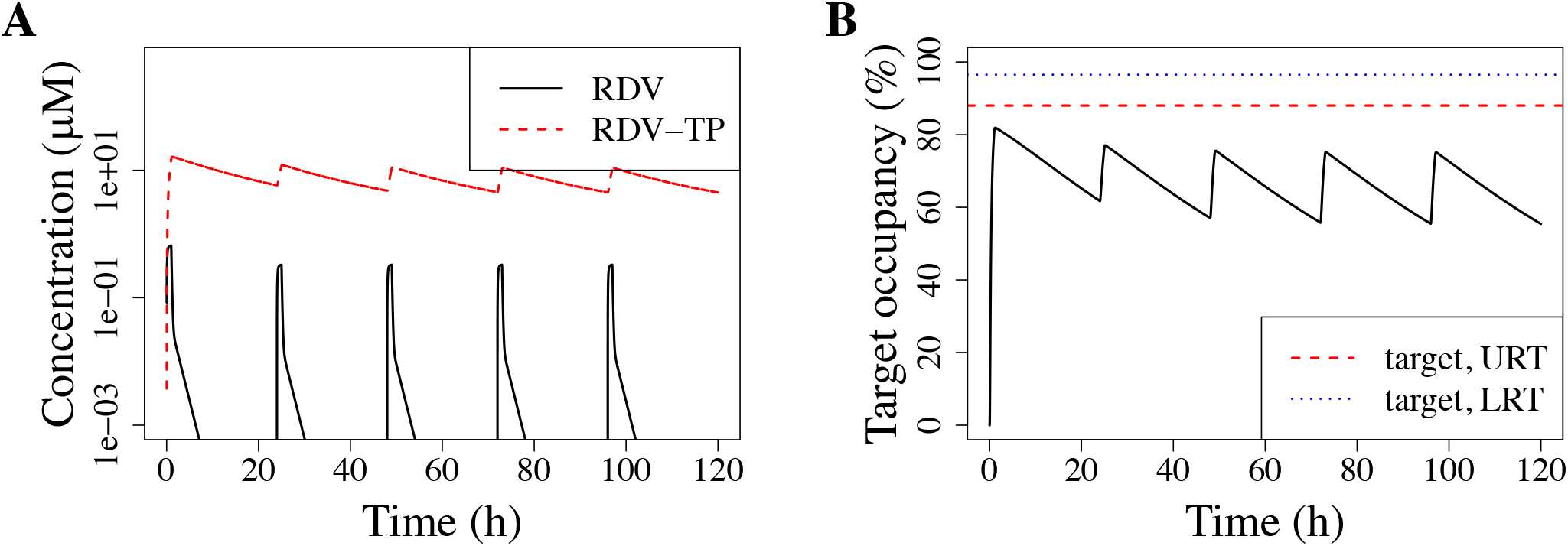
Pharmacokinetics and target occupancy with standard RDV dosing. (**a**) Shows the simulated pharmacokinetics for standard dosing of RDV, i.e. once daily 200 mg loading dose, 100 mg thereafter, infused over 1h (recommended range 30min to 2h). We simulate 5 days of treatment (time given in hours). The concentration of RDV in plasma (black line) and RDV-TP in blood cells (PBMCs, red line), is given in μM. (**b**) Shows the resulting target occupancy as obtained from the intracellular RDV-TP concentration in (A) and equation (2). The red dotted line indicates the minimal effective target occupancy for the LRT and the green dotted line the same for the URT. Both were calculated with equation (3).

Our results thus indicate that the standard dosing regimen of RDV fails to suppress viral replication in either the URT or LRT. In fact, it lowers the viral production rate by an average of ∼70%, where at least 88% would be necessary to suppress viral replication in the URT in an average patient. However, our results also indicate the RDV is not entirely ineffective, it is only dosed suboptimally.

### 3.2 Model-predicted optimal dosing rate

Using our nonlinear model equation (1), we can predict an RDV dosing rate that would approximately maximize the intracellular active metabolite RDV-TP, thereby maximizing efficacy of RDV. RDV data [15] (Figure 1a&b) shows that plasma RDV achieves steady-state rapidly after initiating the infusion, and decays very rapidly after the infusion ends. We therefore use the RDV steady-state from equation (1), 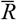, to then maximize the amount of RDV ultimately converted to 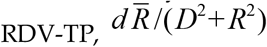, as a function of the dosing rate *r*= *q*_0_ / *τ*. Details are provided in the Supplementary Materials Text S1. Thus we predict with our model that a dosing rate of approximately 168 mg/h maximizes intracellular RDV-TP, thereby maximizing the efficacy of RDV therapy for COVID-19. Thus for example, if we a 1-hour infusion is planned, a total dose of approximately 168 mg should maximize the drug efficacy, while for 2-hour infusion or 30-minute infusions, total doses of approximately 336 mg or 84 mg, respectively, would maximize RDV drug efficacy. In the following, we use this optimal infusion rate (rounding both duration and dosing) to explore the efficacy of several dosing regimens.

### 3.3 Improved dosing with RDV doses that were tested in phase I trials

Our results indicate that RDV is currently underdosed. We therefore set out to investigate the highest daily dose tested in phase I studies, 225 mg, infused at an optimal rate (80 min). Again, we assess a 5 day treatment regimen. We find (Figure 4) that this dosing regimen performs better. However, viral replication would also in this case only be suppressed in the URT at peak concentrations. In the LRT, the necessary concentration would never been reached.

**Figure 4.**
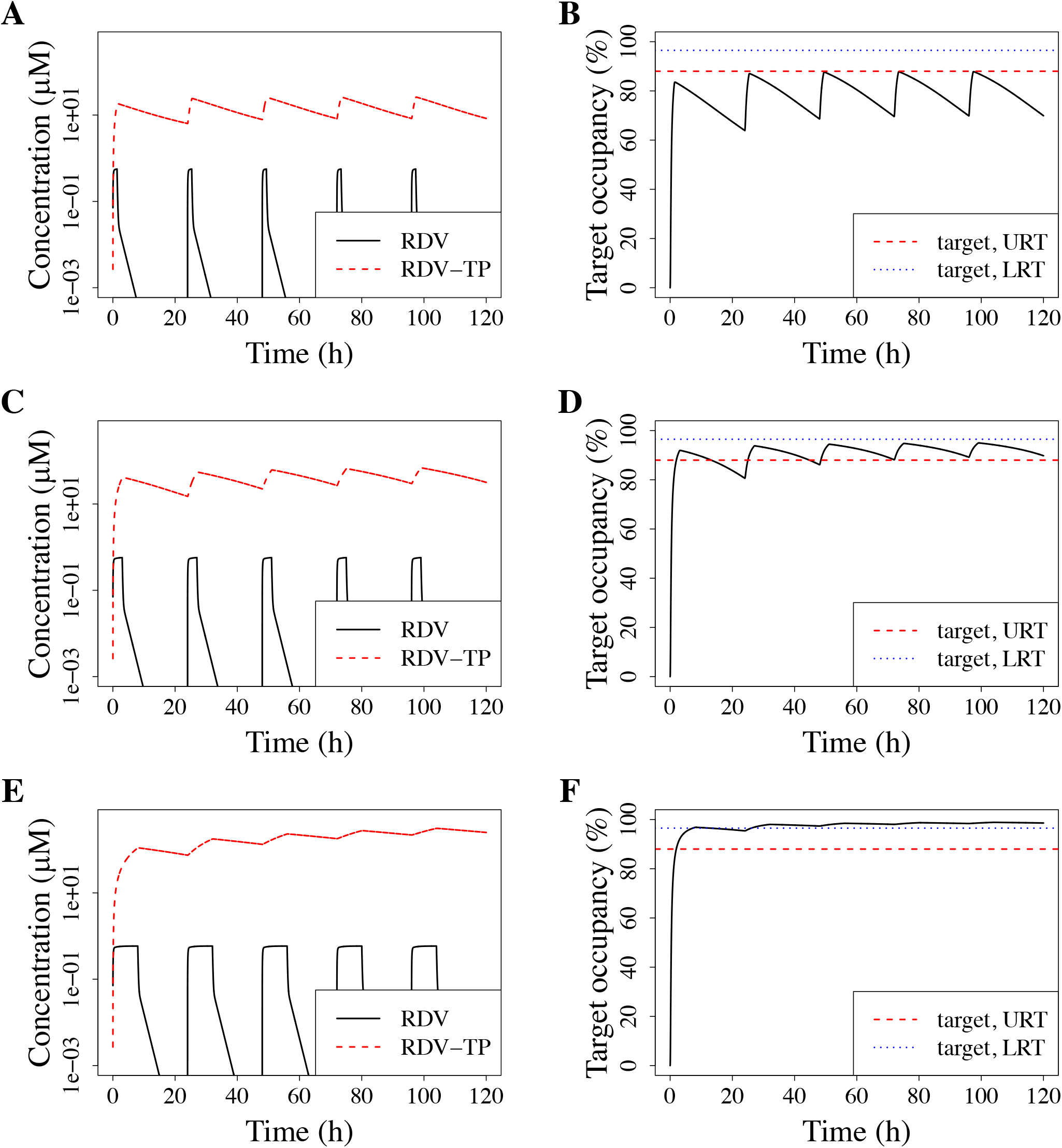
Pharmacokinetics and target occupancy with alternative dosing regimens. (**a**) & (**b**): 225 mg infused over 80 min. (**c**) & (**d**): 500 mg infused over 3 h. (**e**) & (**f**): 1350 mg infused over 8 h. (**a**), (**c**) & (**e**) show the simulated pharmacokinetics. We simulate 5 days of treatment (time given in hours). The concentration of RDV in plasma (black line) and RDV-TP in blood cells (PBMCs, red line), is given in μM. (**b**), (**d**) & (**f**) show the resulting target occupancy as obtained from the intracellular RDV-TP concentration on the left hand panel and equation (2). The red dotted line indicates the minimal effective target occupancy for the LRT and the green dotted line the same for the URT. Both were calculated with equation (3).

### 3.4 Alternative: longer infusion

To the best of our knowledge, it is currently unclear whether RDV toxicity is mediated by RDV itself or any of its metabolites, and whether this toxicity is correlated to peak concentrations, average exposure or time that the concentration exceeds a certain threshold. In addition to hepatotoxicity, side effects that peak during the time of infusion have been described [41]. This might hint that RDV toxicity is driven by peak RDV concentrations in plasma. If this was true, one could keep the peak concentration of RDV in plasma constant but maximize intracellular RDV-TP concentration by extending the infusion time. Here, we investigate which RDV doses infused at an optimal rate would allow viral suppression in the upper and lower respiratory tract, respectively. We find that 500 mg infused over 3 h would suppress viral replication in the upper respiratory tract (Figure 4 c,d,e & f). For suppressing viral replication in the lower respiratory tract, 1350 mg RDV would have to be infused overnight (8 h).

At this time point, it is entirely unclear whether RDV toxicity is indeed mediated by peak plasma concentrations and whether such dosing regimens would be safe. However, our results do indicate that RDV therapy could be further optimized if safe and calls both for dose ranging studies and clarification which metabolites contribute to toxicity.

### 3.5 Avoiding a potential DR mutant

As mentioned in the introduction, a mutation conferring mild resistance (2.5x increased EC_50_) was described emerged in a remdesivir-treated patient [31–33]. This mutation did not carry a detectable fitness cost. If this mutant has no fitness costs in all relevant environments in the host and during transmission, we would expect it to arise quite frequently in remdesivir treated patients and ultimately to spread between patients, irrespective of mitigation strategies. Resistance mutations without fitness costs in vitro have been described in other anti-infectives [42], but frequently they do not spread epidemiologically, hinting at fitness costs in vivo and/or during transmission. Mutations conferring higher levels of resistance to remdesivir but a significant (but unspecified) fitness cost in vitro have been described already before the pandemic in the very closely related MERS virus [43]. Generally, spontaneous resistance mutations can lower the efficacy of antiviral polymerase inhibitors up to 20x [44]. When resistant mutants carry a fitness cost, their spread can more easily be mitigated. A useful concept for this is avoiding drug concentrations in the resistance selection window (see methods section 2.3). The resistance selection window describes antiviral concentrations which are high enough that the resistant virus spreads better within host, but low enough that the resistant virus is not eradicated, i.e. antiviral concentrations that select for resistance. Figure 5 illustrates this concept with arbitrary resistance mutations that carry a 20% fitness cost and have a 5x or 20x reduced susceptibility to remdesivir, either because of reduced drug-target affinity or because of increased excision repair (the latter would increase the drug-target dissociation rate).

**Figure 5.**
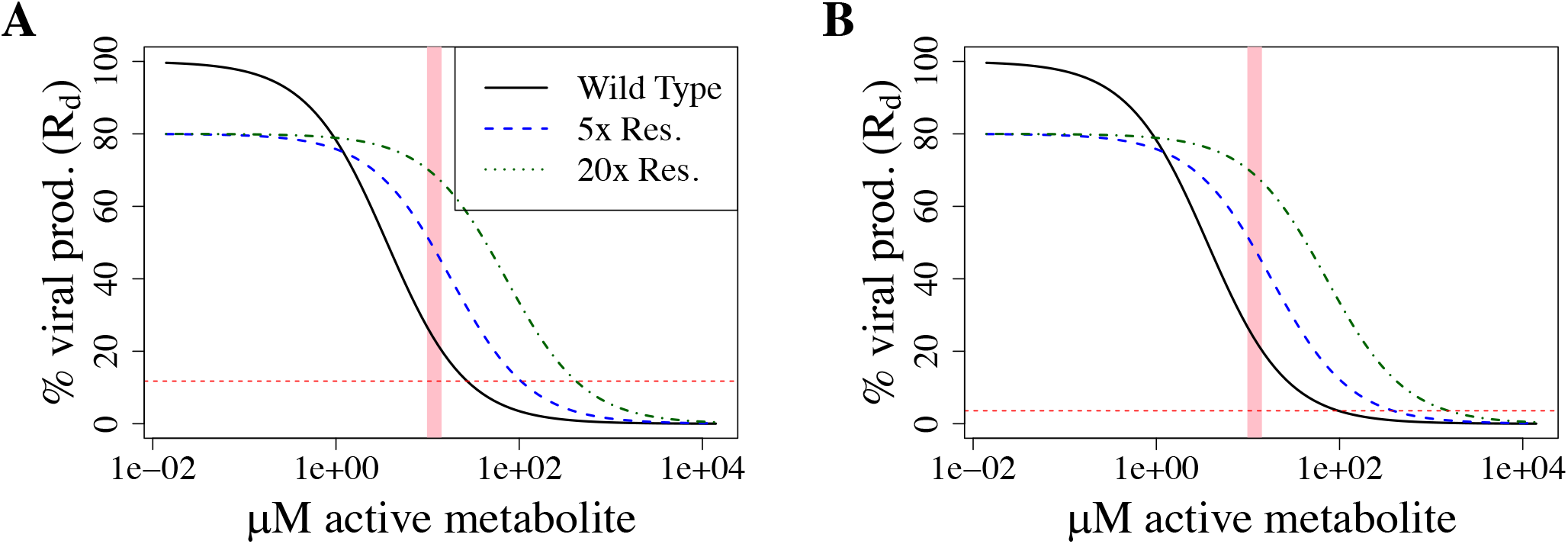
Resistance-selection window for upper and lower respiratory tract. (**a**) Resistance selection window for the upper respiratory tract (R_0_= 8.5), (**b**) resistance selection window for the lower respiratory tract (R_0_= 27.5). In both figures, the x-axis shows the intracellular RDV-TP concentration on the x-axis and the relative viral production rate (and therefore R_d_) on the y-axis (as obtained with equation (3)). The solid black line shows the wild-type, the dotted lines arbitrary resistance mutations with a 20% fitness cost and a 5x (blue) or 20x (green) increased K_D_. The pink shaded area shows the intracellular RDV-TP ranger measured in phase I trials [15]. The red dotted line marks when R_d_ would become 1 for the upper (**a**) and lower (**b**) respiratory tract.

Although there are case reports of resistance, we do not have a good overview of potential RDV-resistant mutations. We therefore first assess the minimal fitness with which newly emerging resistant mutants can be selected for (equation 8) with a range of reduced drug-target binding. Figure 6a illustrates that for within-host R_0_ larger than two, resistance mutations may carry more than 50% fitness costs and yet can be theoretically selected for. This means that there are drug concentrations that are able to select for resistance, even if it is a very narrow concentration range just below the minimal effective concentration for the resistant strain. Importantly, R_0_ is likely much larger than 2, it has been estimated to be 8.5 [4.3-25] for the URT and 27.5 [19.8-42.1] for the LRT [34].

**Figure 6.**
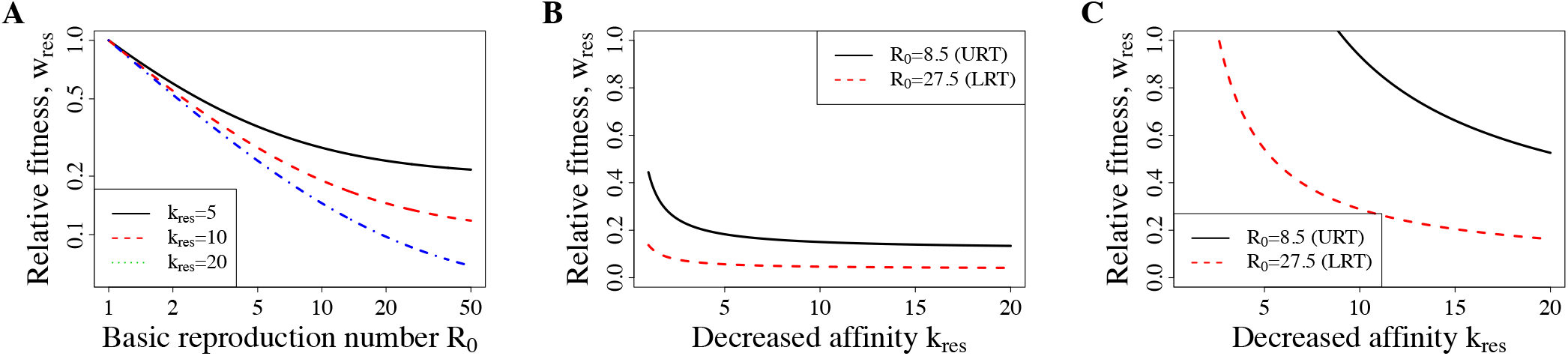
Fitness requirements for selection of resistant mutants. (**a**) Shows the minimal resistance with which newly emerging resistant mutants can be selected for dependent on the baseline R_0_ in a patient, as obtained with equation (8). The x-axis shows R_0_ (estimated between 4.3 and 42.1 depending on body compartment and patient). The y-axis shows the relative fitness costs. (**b**) & (**c**) show the minimal fitness with which newly emerging resistant mutants can be selected for depending on the decrease in drug-target affinity, as obtained with equation (7). The x-axis shows the parameter k_res_, which describes the factor by which drug-target binding is decreased by a resistance mutation, the y-axis the relative fitness. The black line shows results for the upper respiratory tract and the red line for the lower respiratory tract. (**b**) Shows the results for the standard treatment. (**c**) Shows the results for a 1350 mg/8 h infusion.

We then go on to assess the maximal fitness of mutants that could be suppressed with the current dosing regimen that results in an intracellular concentration of approximately 10 µM RDV-TP (Figure 6b&c). We find that for most realistic decreases in binding affinity, the maximal fitness of resistant strains would be below 0.2 in the URT and 0.1 in the LRT. In other words, resistant strains would have to have extremely severe fitness deficiencies for the current dosing to suppress resistance. The highest dose we simulated here, 1350 mg infused overnight (8 h) results in a mean intracellular RDV-TP content of around 250 µM in the second half of the therapy (i.e., between 60-120 h). This would be sufficient to suppress resistance in the URT, but in the LRT mutants with an intermediate fitness and intermediate decreases in drug-target affinity might arise.

## 4. Discussion

In this work, we developed a PK/PD model that allows predicting the treatment efficacy of remdesivir, which is currently the only approved antiviral to treat COVID-19 patients. In contrast to earlier work, our model is built on clinical data from phase I rather than extrapolated to human patients from preclinical murine data. Our model predicts that remdesivir treatment efficacy could be substantially improved by i) optimizing the infusion rate and ii) increasing the overall dose.

Typical pharmacodynamic models rely on fitting empirical sigmoidal dose-response curves to data obtained in vitro. For remdesivir, this approach is difficult, because remdesivir is a prodrug and has to be converted to its active form, RDV-TP upon cell entry [36]. Furthermore, remdesivir is very unstable and rapidly decays at temperatures used in cell culture (37 °C), such that over the time course of an experiment (15-48 h), only a miniscule fraction of the originally used drug concentration is left [38]. Thus, the measured EC_50_s from cell culture have also been termed “pseudo-EC_50_s” [45]. It is difficult to use these pseudo-EC_50_s directly in PK-PD models because remdesivir does not follow the same thermal degradation in vivo (rather, remdesivir decay is described by a pharmacokinetic model) and because diffusion barriers and the necessary metabolic steps of drug activation complicate the relationship between extracellular remdesivir and intracellular RDV-TP. Thus, using only plasma remdesivir levels coupled to this pseudo-EC_50_ will likely give inaccurate results. Therefore, we employ a pharmacokinetic model to predict the intracellular concentration of RDV-TP and then use RDV-TP target engagement calculated based on a predicted affinity constant from structural studies [35]. Although this approach makes multiple assumptions, we find that our model accurately predicts EC_50_s measured in cell culture using extracellularly supplied initial remdesivir concentrations.

However, the EC_50_ alone is not enough to predict optimal therapy, since it might be necessary to achieve very high levels (not only half-maximal) to suppress viral spread within host. A typical target would be achieving 90% viral suppression (EC_90_). Here, we use the speed with which SARS-CoV-2 spreads through the body (within-host R_0_) to determine how much viral replication has to be suppressed to clear the virus. While we expect that the EC_90_ is sufficient in the upper respiratory tract where the virus spreads less quickly, we expect that 96.5% target binding and therefore viral suppression are needed to inhibit viral replication in the lower respiratory tract.

Based on our pharmacokinetic model, which is calibrated to RDV-TP concentrations in blood (PBMC) and not lung tissue, we expect that the current standard dosing regimen fails to suppress viral replication in both the upper and the lower respiratory tract. Lung tissue concentrations are likely even lower, such that our estimates are conservative [25].

Nevertheless, our results do not indicate that remdesivir is ineffective, they indicate that the current dosing has room for improvement if toxicity allows. This is in line with clinical trials that report a shorter time to recovery [1,46], newer meta-analyses that support a slightly reduced mortality [7] and recent findings that the viral load declines more quickly in patients receiving remdesivir [11]. We therefore would like to argue for more dose-ranging and dose-fractionation trials. To minimize toxicity, it would also be important to understand whether remdesvir itself or any of its metabolites drive toxicity, and whether this toxicity correlates best with peak concentrations (C_max_) or average exposure (AUC).

Remdesivir resistance mutations have been characterized both clinically and experimentally. Unfortunately, one mutation conferring mild resistance does not carry any fitness cost in vitro [31–33]. Moreover, our model predicts that the current standard regimen strongly selects for resistance to remdesivir for a vast majority of the credible parameter space. We would therefore argue for increasing efforts to find potential resistance mutations in patients treated with remdesivir. Combining remdesivir with one or more other antivirals would not only improve outcome, it likely would also reduce resistance evolution [19–23]. This might be especially important if remdesivir is used as a template to design new compounds with increased efficacy [47]. Resistance against older, less effective anti-infectives often provides a stepping stone to resistance against improved but similar compounds [48].

Taken together, our work indicates that remdesivir is expected to be mildly effective and therefore a useful companion drug in COVID-19 treatment. However, dosing should be improved and/or antiviral companion drugs should be employed to safeguard against resistance.

## Data Availability

Data employed in this study was previously published. The authors of this manuscript were not involved in the studies from which the data derived. Those studies are cited in the manuscript.

## Supplementary Materials

The following are available online: Table S1, Pharmacokinetic parameter estimates for metabolite RDV-TP from single-dose experiments, with different RDV dose size and infusion durations, compared to our pharmacokinetic model prediction. Figure S1, Median RDV plasma concentration from single-dose experiments, with different RDV dose size and infusion duration, compared to our pharmacokinetic model prediction. Figure S2, Extracellular RDV concentrations and intracellular RDV-TP concentrations during in vitro dose-response experiments. Finally, Text S1, Details on development of pharmacokinetic model describing RDV and RDV-TP dynamics in plasma and associated parameter estimation, with the optimal dosing approximation calculation.

## Author Contributions

Conceptualization, JMC and PAzW.; methodology, JMC and PAzW.; software, JMC and PAzW.; validation, JMC and PAzW.; formal analysis, JMC and PAzW.; investigation, JMC and PAzW.; resources, JMC and PAzW.; data curation, JMC and PAzW.; writing—original draft preparation, JMC and PAzW.; writing—review and editing, JMC and PAzW.; visualization JMC and PAzW.; supervision, NA.; project administration, JMC and PAzW.; funding acquisition, JMC and PAzW All authors have read and agreed to the published version of the manuscript.

## Institutional Review Board Statement

Not applicable.

## Informed Consent Statement

Not applicable.

## Data Availability Statement

Not applicable.

## Acknowledgments

We would like to thank Sören Abel, Fabrizio Clarelli, Jingyi Liang and Colin Hemez for discussions and input.

## References

1. Beigel, J.H.; Tomashek, K.M.; Dodd, L.E.; Mehta, A.K.; Zingman, B.S.; Kalil, A.C.; Hohmann, E.; Chu, H.Y.; Luetkemeyer, A.; Kline, S.; et al. Remdesivir for the Treatment of Covid-19 — Final Report. N. Engl. J. Med. 2020, 383, 1813–1826, doi:10.1056/NEJMoa2007764.

2. First COVID-19 Treatment Recommended for EU Authorisation.

3. FDA Approves First Treatment for COVID-19.

4. de Wit, E.; Feldmann, F.; Cronin, J.; Jordan, R.; Okumura, A.; Thomas, T.; Scott, D.; Cihlar, T.; Feldmann, H. Prophylactic and Therapeutic Remdesivir (GS-5734) Treatment in the Rhesus Macaque Model of MERS-CoV Infection. Proc. Natl. Acad. Sci. 2020, 117, 6771–6776, doi:10.1073/pnas.1922083117.

5. NIH Clinical Trial Shows Remdesivir Accelerates Recovery from Advanced COVID-19.

6. WHO Solidarity Trial Consortium Repurposed Antiviral Drugs for Covid-19 — Interim WHO Solidarity Trial Results. N. Engl. J. Med. 2021, 384, 497–511, doi:10.1056/NEJMoa2023184.

7. Kaka, A.S.; MacDonald, R.; Greer, N.; Vela, K.; Duan-Porter, W.; Obley, A.; Wilt, T.J. Major Update: Remdesivir for Adults With COVID-19: A Living Systematic Review and Meta-Analysis for the American College of Physicians Practice Points. Ann. Intern. Med. 2021, 174, 663–672, doi:10.7326/M20-8148.

8. Garibaldi, B.T.; Fiksel, J.; Muschelli, J.; Robinson, M.L.; Rouhizadeh, M.; Perin, J.; Schumock, G.; Nagy, P.; Gray, J.H.; Malapati, H.; et al. Patient Trajectories Among Persons Hospitalized for COVID-19: A Cohort Study. Ann. Intern. Med. 2021, 174, 33–41, doi:10.7326/M20-3905.

9. Néant, N.; Lingas, G.; Le Hingrat, Q.; Ghosn, J.; Engelmann, I.; Lepiller, Q.; Gaymard, A.; Ferré, V.; Hartard, C.; Plantier, J.-C.; et al. Modeling SARS-CoV-2 Viral Kinetics and Association with Mortality in Hospitalized Patients from the French COVID Cohort. Proc. Natl. Acad. Sci. 2021, 118, e2017962118, doi:10.1073/pnas.2017962118.

10. Goyal, A.; Duke, E.R.; Cardozo-Ojeda, E.F.; Schiffer, J.T. Mathematical Modeling Explains Differential SARS CoV-2 Kinetics in Lung and Nasal Passages in Remdesivir Treated Rhesus Macaques. bioRxiv 2020, 2020.06.21.163550, doi:10.1101/2020.06.21.163550.

11. Regan, J.; Flynn, J.P.; Rosenthal, A.; Jordan, H.; Li, Y.; Chishti, R.; Giguel, F.; Corry, H.; Coxen, K.; Fajnzylber, J.; et al. Viral Load Kinetics of SARS-CoV-2 In Hospitalized Individuals with COVID-19. Open Forum Infect. Dis. 2021, doi:10.1093/ofid/ofab153.

12. Monteil, V.; Dyczynski, M.; Lauschke, V.M.; Kwon, H.; Wirnsberger, G.; Youhanna, S.; Zhang, H.; Slutsky, A.S.; Hurtado del Pozo, C.; Horn, M.; et al. Human Soluble ACE2 Improves the Effect of Remdesivir in SARS-CoV-2 Infection. EMBO Mol. Med. 2021, 13, doi:10.15252/emmm.202013426.

13. Kalil, A.C.; Patterson, T.F.; Mehta, A.K.; Tomashek, K.M.; Wolfe, C.R.; Ghazaryan, V.; Marconi, V.C.; Ruiz-Palacios, G.M.; Hsieh, L.; Kline, S.; et al. Baricitinib plus Remdesivir for Hospitalized Adults with Covid-19. N. Engl. J. Med. 2021, 384, 795–807, doi:10.1056/NEJMoa2031994.

14. Adaptive COVID-19 Treatment Trial 4 (ACTT-4).

15. Humeniuk, R.; Mathias, A.; Cao, H.; Osinusi, A.; Shen, G.; Chng, E.; Ling, J.; Vu, A.; German, P. Safety, Tolerability, and Pharmacokinetics of Remdesivir, An Antiviral for Treatment of COVID-19, in Healthy Subjects. Clin. Transl. Sci. 2020, doi:10.1111/cts.12840.

16. Eastman, R.T.; Roth, J.S.; Brimacombe, K.R.; Simeonov, A.; Shen, M.; Patnaik, S.; Hall, M.D. Remdesivir: A Review of Its Discovery and Development Leading to Emergency Use Authorization for Treatment of COVID-19. ACS Cent. Sci. 2020, 6, 672–683, doi:10.1021/acscentsci.0c00489.

17. Humeniuk, R.; Mathias, A.; Kirby, B.J.; Lutz, J.D.; Cao, H.; Osinusi, A.; Babusis, D.; Porter, D.; Wei, X.; Ling, J.; et al. Pharmacokinetic, Pharmacodynamic, and Drug-Interaction Profile of Remdesivir, a SARS-CoV-2 Replication Inhibitor. Clin. Pharmacokinet. 2021, 60, 569–583, doi:10.1007/s40262-021-00984-5.

18. Xu, Y.; Barauskas, O.; Kim, C.; Babusis, D.; Murakami, E.; Kornyeyev, D.; Lee, G.; Stepan, G.; Perron, M.; Bannister, R.; et al. Off-Target In Vitro Profiling Demonstrates That Remdesivir Is a Highly Selective Antiviral Agent. Antimicrob. Agents Chemother. 2021, 65, doi:10.1128/AAC.02237-20.

19. Mager, D.E.; Jusko, W.J. General Pharmacokinetic Model for Drugs Exhibiting Target-Mediated Drug Disposition. J. Pharmacokinet. Pharmacodyn. 2001, 28, 507–532, doi:10.1023/a:1014414520282.

20. Shen, L.; Rabi, S.A.; Sedaghat, A.R.; Shan, L.; Lai, J.; Xing, S.; Siliciano, R.F. A Critical Subset Model Provides a Conceptual Basis for the High Antiviral Activity of Major HIV Drugs. Sci. Transl. Med. 2011, 3, 91ra63–91ra63, doi:10.1126/scitranslmed.3002304.

21. Walkup, G.K.; You, Z.; Ross, P.L.; Allen, E.K.H.; Daryaee, F.; Hale, M.R.; O’Donnell, J.; Ehmann, D.E.; Schuck, V.J.A.; Buurman, E.T.; et al. Translating Slow-Binding Inhibition Kinetics into Cellular and in Vivo Effects. Nat. Chem. Biol. 2015, 11, 416–423, doi:10.1038/nchembio.1796.

22. Abel zur Wiesch, P.; Abel, S.; Gkotzis, S.; Ocampo, P.; Engelstädter, J.; Hinkley, T.; Magnus, C.; Waldor, M.K.; Udekwu, K.; Cohen, T. Classic Reaction Kinetics Can Explain Complex Patterns of Antibiotic Action. Sci. Transl. Med. 2015, 7, 287ra73–287ra73, doi:10.1126/scitranslmed.aaa8760.

23. Baeder, D.Y.; Yu, G.; Hozé, N.; Rolff, J.; Regoes, R.R. Antimicrobial Combinations: Bliss Independence and Loewe Additivity Derived from Mechanistic Multi-Hit Models. Philos. Trans. R. Soc. B Biol. Sci. 2016, 371, 20150294, doi:10.1098/rstb.2015.0294.

24. Wang, W.; Wang, X.; Doddareddy, R.; Fink, D.; McIntosh, T.; Davis, H.M.; Zhou, H. Mechanistic Pharmacokinetic/Target Engagement/Pharmacodynamic (PK/TE/PD) Modeling in Deciphering Interplay Between a Monoclonal Antibody and Its Soluble Target in Cynomolgus Monkeys. AAPS J. 2014, 16, 129–139, doi:10.1208/s12248-013-9545-8.

25. Hanafin, P.O.; Jermain, B.; Hickey, A.J.; Kabanov, A.V.; Kashuba, A.D.; Sheahan, T.P.; Rao, G.G. A Mechanism-based Pharmacokinetic Model of Remdesivir Leveraging Interspecies Scaling to Simulate COVID-19 Treatment in Humans. CPT Pharmacomet. Syst. Pharmacol. 2021, 10, 89–99, doi:10.1002/psp4.12584.

26. Nair, A.; Jacob, S. A Simple Practice Guide for Dose Conversion between Animals and Human. J. Basic Clin. Pharm. 2016, 7, 27, doi:10.4103/0976-0105.177703.

27. Leenaars, C.H.C.; Kouwenaar, C.; Stafleu, F.R.; Bleich, A.; Ritskes-Hoitinga, M.; De Vries, R.B.M.; Meijboom, F.L.B. Animal to Human Translation: A Systematic Scoping Review of Reported Concordance Rates. J. Transl. Med. 2019, 17, doi:10.1186/s12967-019-1976-2.

28. Pennings, P.S. HIV Drug Resistance: Problems and Perspectives. Infect. Dis. Rep. 2013, 5, e5, doi:10.4081/idr.2013.s1.e5.

29. Lampejo, T. Influenza and Antiviral Resistance: An Overview. Eur. J. Clin. Microbiol. Infect. Dis. 2020, 39, 1201–1208, doi:10.1007/s10096-020-03840-9.

30. Martinot, M.; Jary, A.; Fafi-Kremer, S.; Leducq, V.; Delagreverie, H.; Garnier, M.; Pacanowski, J.; Mékinian, A.; Pirenne, F.; Tiberghien, P.; et al. Remdesivir Failure with SARS-CoV-2 RNA-Dependent RNA-Polymerase Mutation in a B-Cell Immunodeficient Patient with Protracted Covid-19. Clin. Infect. Dis. 2020, doi:10.1093/cid/ciaa1474.

31. Szemiel, A.M.; Merits, A.; Orton, R.J.; MacLean, O.; Pinto, R.M.; Wickenhagen, A.; Lieber, G.; Turnbull, M.L.; Wang, S.; Mair, D.; et al. <em>In Vitro</Em> Evolution of Remdesivir Resistance Reveals Genome Plasticity of SARS-CoV-2. bioRxiv 2021, 2021.02.01.429199, doi:10.1101/2021.02.01.429199.

32. Padhi, A.K.; Shukla, R.; Saudagar, P.; Tripathi, T. High-Throughput Rational Design of the Remdesivir Binding Site in the RdRp of SARS-CoV-2: Implications for Potential Resistance. iScience 2021, 24, 101992, doi:10.1016/j.isci.2020.101992.

33. Mari, A.; Roloff, T.; Stange, M.; Søgaard, K.K.; Asllanaj, E.; Tauriello, G.; Alexander, L.T.; Schweitzer, M.; Leuzinger, K.; Gensch, A.; et al. Global Surveillance of Potential Antiviral Drug Resistance in SARS-CoV-2: Proof of Concept Focussing on the RNA-Dependent RNA Polymerase. medRxiv 2021, 2020.12.28.20248663, doi:10.1101/2020.12.28.20248663.

34. Ke, R.; Zitzmann, C.; Ribeiro, R.M.; Perelson, A.S. Kinetics of SARS-CoV-2 Infection in the Human Upper and Lower Respiratory Tracts and Their Relationship with Infectiousness. medRxiv 2020, 2020.09.25.20201772, doi:10.1101/2020.09.25.20201772.

35. Zhang, L.; Zhou, R. Structural Basis of the Potential Binding Mechanism of Remdesivir to SARS-CoV-2 RNA-Dependent RNA Polymerase. J. Phys. Chem. B 2020, 124, 6955–6962, doi:10.1021/acs.jpcb.0c04198.

36. Götte, M. Remdesivir for the Treatment of Covid-19: The Value of Biochemical Studies. Curr. Opin. Virol. 2021, 49, 81– 85, doi:10.1016/j.coviro.2021.04.014.

37. Perelson, A.S. Modelling Viral and Immune System Dynamics. Nat. Rev. Immunol. 2002, 2, 28–36, doi:10.1038/nri700.

38. Avataneo, V.; de Nicolò, A.; Cusato, J.; Antonucci, M.; Manca, A.; Palermiti, A.; Waitt, C.; Walimbwa, S.; Lamorde, M.; di Perri, G.; et al. Development and Validation of a UHPLC-MS/MS Method for Quantification of the Prodrug Remdesivir and Its Metabolite GS-441524: A Tool for Clinical Pharmacokinetics of SARS-CoV-2/COVID-19 and Ebola Virus Disease. J. Antimicrob. Chemother. 2020, 75, 1772–1777, doi:10.1093/jac/dkaa152.

39. Pruijssers, A.J.; George, A.S.; Schäfer, A.; Leist, S.R.; Gralinksi, L.E.; Dinnon, K.H.; Yount, B.L.; Agostini, M.L.; Stevens, L.J.; Chappell, J.D.; et al. Remdesivir Inhibits SARS-CoV-2 in Human Lung Cells and Chimeric SARS-CoV Expressing the SARS-CoV-2 RNA Polymerase in Mice. Cell Rep. 2020, 32, 107940, doi:10.1016/j.celrep.2020.107940.

40. Wang, M.; Cao, R.; Zhang, L.; Yang, X.; Liu, J.; Xu, M.; Shi, Z.; Hu, Z.; Zhong, W.; Xiao, G. Remdesivir and Chloroquine Effectively Inhibit the Recently Emerged Novel Coronavirus (2019-nCoV) in Vitro. Cell Res. 2020, 30, 269–271, doi:10.1038/s41422-020-0282-0.

41. Fan, Q.; Zhang, B.; Ma, J.; Zhang, S. Safety Profile of the Antiviral Drug Remdesivir: An Update. Biomed. Pharmacother. 2020, 130, 110532, doi:10.1016/j.biopha.2020.110532.

42. Baker, S.; Duy, P.T.; Nga, T.V.T.; Dung, T.T.N.; Phat, V.V.; Chau, T.T.; Turner, A.K.; Farrar, J.; Boni, M.F. Fitness Benefits in Fluoroquinolone-Resistant Salmonella Typhi in the Absence of Antimicrobial Pressure. eLife 2013, 2, doi:10.7554/eLife.01229.

43. Agostini, M.L.; Andres, E.L.; Sims, A.C.; Graham, R.L.; Sheahan, T.P.; Lu, X.; Smith, E.C.; Case, J.B.; Feng, J.Y.; Jordan, R.; et al. Coronavirus Susceptibility to the Antiviral Remdesivir (GS-5734) Is Mediated by the Viral Polymerase and the Proofreading Exoribonuclease. mBio 2018, 9, doi:10.1128/mBio.00221-18.

44. Mo, H.; Lu, L.; Pilot-Matias, T.; Pithawalla, R.; Mondal, R.; Masse, S.; Dekhtyar, T.; Ng, T.; Koev, G.; Stoll, V.; et al. Mutations Conferring Resistance to a Hepatitis C Virus (HCV) RNA-Dependent RNA Polymerase Inhibitor Alone or in Combination with an HCV Serine Protease Inhibitor in Vitro. Antimicrob. Agents Chemother. 2005, 49, 4305–4314, doi:10.1128/AAC.49.10.4305-4314.2005.

45. Shi, J.; Xiao, Y.; Zhang, Y.; Geng, D.; Cong, D.; Shi, K.X.; Knapp, R.J. Challenges of Drug Development during the COVID-19 Pandemic: Key Considerations for Clinical Trial Designs. Br. J. Clin. Pharmacol. 2021, 87, 2170–2185, doi:10.1111/bcp.14629.

46. Lai, C.-C.; Chen, C.-H.; Wang, C.-Y.; Chen, K.-H.; Wang, Y.-H.; Hsueh, P.-R. Clinical Efficacy and Safety of Remdesivir in Patients with COVID-19: A Systematic Review and Network Meta-Analysis of Randomized Controlled Trials. J. Antimicrob. Chemother. 2021, doi:10.1093/jac/dkab093.

47. Gleeson, M.P.; Hersey, A.; Montanari, D.; Overington, J. Probing the Links between in Vitro Potency, ADMET and Physicochemical Parameters. Nat. Rev. Drug Discov. 2011, 10, 197–208, doi:10.1038/nrd3367.

48. Wang, Y.C.; Lipsitch, M. Upgrading Antibiotic Use within a Class: Tradeoff between Resistance and Treatment Success. Proc. Natl. Acad. Sci. 2006, 103, 9655–9660, doi:10.1073/pnas.0600636103.

