## Supplementary Materials Text 1 for "Remdesivir to treat COVID-19: can dosing be optimized?"

### Remdesivir to treat COVID-19: can dosing be optimized?
### -- Supporting Methods –

Jessica M. Conway and Pia Abel zur Wiesch

#### 1 Pharmacokinetic model and parameter estimation

##### Model of remdesivir prodrug in plasma

We begin by estimating model parameters for remdesivir in plasma only. We consider extra-cellular concentration of RDV in plasma and the periphery, $R$ and $P$.

$$\begin{aligned}\frac{dR}{dt} &= \frac{q_0}{V_r}H(t-\tau) - (\beta+\delta_R)R + kP \\ \frac{dP}{dt} &= \beta R - (k+\delta_P)P\end{aligned} \tag{1}$$

Note that we assume for this model a single dose, initiated at time 0, with duration $\tau$, to compare with data in Humeniuk et al. (2020) [1]. We later append to this model a equation describing the active triphosphate metabolite, RDV-TP (GS-443902).

We fit this model to the longitudinal data in Figures 1a and 2a of Humeniuk et al. (2020) [1]. Specifically we fit the model to the median values. Since we cannot extract the error bars from the graphs (they overlap), we cannot weight the fitting algorithm appropriately. This poses a difficulty because the dynamics of 75mg over 2 hours (Fig 1a) don't follow the trends set by the other dosings, but the data points in the tail, when drug concentrations decay, have large or no error bars. We therefore fit the model including and excluding this data set. However, we keep it in our analysis of the fit for transparency (Supporting Figure A1). Note that the model explain the 75mg dosing data when drug concentrations are high, when the majority of the conversion to intracellular metabolite takes place (see below), and fails to explain reported median drug concentrations only when drug concentrations are low. For fitting, we minimize the sum-of-square difference between the model and the median RDV concentration values for 3mg, 10mg, 30mg, 150mg, and 225mg infused over two hours, and 150mg infused over 1 hour, using Optim in R, with the Nelder-Mead algorithm followed up by stochastic annealing (SANN) to increase confidence that we have a likely global minimum. Parameter estimates are given in Table 1.

Table 1: RDV model parameter descriptions.

| Parameter | Description | Units | Estimate |
|---|---|---|---|
| $q_0$ | Total mass of RDV infused | mg | n/a |
| $\tau$ | Duration of infusion | h | n/a |
| $V_r$ | Apparent volume for RDV in plasma | L | 6.09 |
| $\beta$ | Plasma-to-periphery RDV concentration transition rate | $h^{-1}$ | 4.42 |
| $\delta_R$ | Elimination rate in plasma | $h^{-1}$ | 4.16 |
| $k$ | Periphery-to-plasma RDV concentration transition rate | $h^{-1}$ | 0.13 |
| $\delta_P$ | Elimination rate in periphery | $h^{-1}$ | 0.61 |

To demonstrate the goodness-of-fit, in Fig. 1 we show the model predictions plotted against the observations and show the linear regression. With slope and $R^2$ close to 1, we show excellent fit..

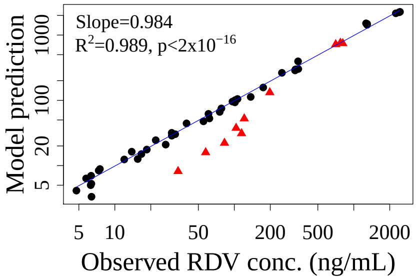


Figure 1: Model predictions vs observations on RDV concentrations for our model eq. (1) for parameter estimates The 75mg data/prediction pairs are represented by the red triangles to show their outlier behavior. We show the linear regression between our model predictions and the data to increase confidence in our model. Note that the relationship is well explained by a linear relationship, with p-values$<10^{-16}$ in both cases.

#### Model extension to predict dynamics of RDV-TP

The exact metabolic pathway that takes plasma RDV to the intracellular active triphosphate RDV-TP (GS-443902) is not yet confirmed [2, 1, 3]. However, there is consensus that intracellular RDV-TP is the active form of the drug [2, 1, 3]. We therefore model RDV and the RDV-TP only. The extended model is

$$\begin{aligned}
\frac{dR}{dt} &= \frac{q_0}{V_r}H(t-\tau) - (\beta+\delta_R)R + kP - d\frac{R^2}{D^2+R^2} \\
\frac{dP}{dt} &= \beta R - (k+\delta_P)P \\
\frac{dA}{dt} &= \sigma d\frac{R^2}{D^2+R^2} - \mu_1 A - \mu_2\frac{A^2}{M^2+A^2}
\end{aligned} \tag{2}$$

Here $R$ represents plasma concentration of RDV, $P$ concentration in the periphery, and $A$ the intracellular RDV-TP concentration. In developing this model, we tested a variety of linear and nonlinear forms and alternative Hill parameters, finding this to best represent dynamics.

To our knowledge longitudinal data on RDV-TP is not available. To estimate parameters, we use parmacokinetic data provided Humeniuk et al. (2020) [1]. Instead of mixing data types for RDV and its active metabolites, we take parameters for RDV as given in Table 1, assuming that the amount that ultimately gets converted to the metabolite is negligible. We will verify this assumption. We then estimate remaining parameters using pharmacokinetic data in Table 4 of Humeniuk et al. (2020) [1], estimating on $C_{max}$, $C_{24}$, $AUC_\infty$, and $t_{1/2}$ for intracellular RDV-TP following a 75 mg infusion over 30 minutes or 2 hours, or a 150 mg infusion over 2 hours. To generate a model-predicted $AUC_\infty$, we integrate our result to 1000 days. To generate a model-predicted "terminal" $t_{1/2}$, we use the decay rate around $t = 48$ hours.

To estimate model parameters $\vec{\theta}$, we seek to minimize the error

$$E(\vec{\theta}) = \sum_{j=1}^{3}\left[\frac{\left(\log(C_{max}) - \log(C_{max}^{pred})\right)^2}{IQR_{C_{max}}/\text{median}_{C_{max}}} + \frac{\left(\log(C_{24}) - \log(C_{24}^{pred})\right)^2}{IQR_{C_{24}}/\text{median}_{C_{24}}} + \frac{\left(\log(AUC_\infty) - \log(AUC_\infty^{pred})\right)^2}{IQR_{AUC_\infty}/\text{median}_{AUC_\infty}} + \frac{\left(\log(t_{1/2}) - \log(t_{1/2}^{pred})\right)^2}{IQR_{t_{1/2}}/\text{median}_{t_{1/2}}}\right]$$

where $j = 1,2,3$ correspond to dosings 75mg over 2 hours, 150mg over 2 hours, and 75 mg over 30 min, respectively. We use the log to balance the values, AUCs are so large they would otherwise dominate. Here we have used weights according to the ratio of the interquartile range (IQR) to the median as described in Arachchige et al. (2020) [4]. We

Table 3: Model predictions vs data on metabolite (Table 4 in Humeniuk et al.). The ** indicate model predictions outside the data's %CV or IQR.

| Dosing | Quantity of interest | Data (%CV or IQR) | Model prediction |
|---|---|---|---|
| 75 mg over 2 hours | $C_{max}$ | 2.5 (16.2) | 2.5 |
| | $C_{24}$ | 2.2 (23.3) | 1.67 |
| | $AUC_\infty$ | 176 (31.1) | 184 |
| | $t_{1/2}$ | 42.7 (30.6-47.4) | 48** |
| 150 mg over 2 hours | $C_{max}$ | 6.0 (46.1) | 8.79 |
| | $C_{24}$ | 3.7 (40.9) | 4.04 |
| | $AUC_\infty$ | 297 (28.3) | 381 |
| | $t_{1/2}$ | 36.0 (27.3-41.5) | 34.4 |
| 75 mg over 30 min | $C_{max}$ | 5.9 (37.7) | 5.47 |
| | $C_{24}$ | 3.3 (55.7) | 2.87 |
| | $AUC_\infty$ | 394 (49.9) | 291 |
| | $t_{1/2}$ | 49.0 (26.6-69.5) | 39.4 |

use the IQR rather than the percent coefficient of variation (%CV) since the %CV is not provided for $t_{1/2}$ and when we attempt to derive one from the provided IQR assuming a normal distribution, we find that the normal distribution poorly explains the $t_{1/2}$. We estimate the IQR and median from the mean and %CV for $C_{max}$, $C_{24}$, $AUC_\infty$, assuming a normal distribution.

To estimate parameters we minimize the eror using Optim in R, with the Nelder-Mead algorithm followed up by stochastic annealing (SANN) to increase confidence that we have a likely global minimum. Parameter estimates are available in Table 2.

Table 2: Metabolite model parameter descriptions.

| **Parameter** | **Description** | **Units** | **Estimate** |
|---|---|---|---|
| $d$ | Nonlinear plasma RDV to intracellular active triphosphate metabolite transport and activation rate | mg/L/h | 31.57 |
| $D$ | RDV concentration yielding 50% of the maximal transport/activation rate | mg/L | 3516.27 |
| $\sigma$ | Conversion of concentrations from mg/L to M (not estimated) | mol/mg | 1 |
| $\mu_1$ | Linear elimination rate of RDV-TP | $h^{-1}$ | 1.55 |
| $M$ | RDV-TP concentration at which the nonlinear elimination rate is 50% its maximum | $\mu M$ | 17.19 |
| $\mu_2$ | Nonlinear metabolite clearance rate | $\mu$M/h | 0.0084 |

We use our parameter estimates to predict $C_{max}$, $C_{24}$, $AUC_\infty$, and $t_{1/2}$ for drug regimen in Table 4 of Humeniuk et al. [1], and compare with the data, using model eq. (2) with parameter estimates deriving from either the weighted or unweighted fitting, given in Table 2. The results are shown in Table 3. All predictions but one land within the %CV of IQR of the data. The exception is the predicted half-life in the 75mg/2 hour dosing. In that case, the predicted half-life is just a hair outside the IQR: the prediction is 48 hours, the IQR is 30.6-47.4 hours. Note also that the model predictions lie within the %CV of reported metabolite $C_{max}$, $C_{24}$, $AUC_{24}$, in Table 16 of the EMA report (200mg dosing, we assume over 2 hours) (not shown).

Finally we verify that our model still predicts RDV dynamics consistent with data, that is, verify that our assumption that the conversion to RDV-TP is negligible relative to prodrug concentrations. Figure 1 and Appendix Figure A1 shows the full model predictions of RDV concentrations. We compare Fig. 1, showing model prediction vs data without active metabolite eq. (1), with Figure 1C/Appendix Figure A1H**,** showing the same in the model with active metabolite eq. (2) and found little change.

#### 2 Predicted optimal remdesivir dosing calculation

Using our nonlinear model eq. (2), we can predict an RDV dosing rate that would maximize the intracellular active metabolite RDV-TP, thereby maximizing efficacy of RDV. Since the model eq. (2) is nonlinear, the answer is not to simply maximize the prodrug plasma concentration.

We note from the data [1] (main text Fig. 1, supporting Fig. 1A) that plasma RDV achieves steady-state rapidly after initiating the infusion. We can compute that that steady-state, from our RDV model assuming constant drug infusion by solving simultaneously the equations

$$0 = \frac{r}{V_r} - (\beta + \delta_R)\bar{R} + k\bar{P} - d\frac{\bar{R}^2}{D^2 + \bar{R}^2} \tag{3}$$

$$0 = \beta\bar{R} - (k + \delta_P)\bar{P} \tag{4}$$

where is the dosing rate $r = q_0/\tau$. We aim to maximize the amount of remdesivir ultimately converted to RDV-TP, which we approximate as the product of the conversion rate at the steady state $\bar{R}$, $d\bar{R}/(D^2 + \bar{R}^2)$, and the infusion duration $\tau$. We are neglecting here RDV dynamics following end of infusion, but we observe from the data and our model prediction [1] (Fig. 1, Supporting Fig. A1) that that quantity is orders of magnitude smaller. That is, we maximize $d\bar{R}/(D^2 + \bar{R}^2)$, where $\bar{R}$ is the solution of eqs. (3,4), with respect to dosing rate $r$.

Thus we estimate that $\approx$ 168 mg per hour maximizes the intracellular active metabolite. That is, if we plan a 1 hour infusion, a total dose of approximately 168 mg should maximize the drug efficacy; for a 2 hour infusion, a total dose of approximately 336 mg maximizes the drug efficacy; and over half an hour, a total dose of approximately 84 mg would achieve the same aim.

#### References


[1] R. Humeniuk, A. Mathias, H. Cao, A. Osinusi, G. Shen, E. Chng, J. Ling, A. Vu, and P. German. Safety, tolerability, and pharmacokinetics of remdesivir, an antiviral for treatment of COVID-19, in healthy subjects. *Clin Transl Sci*, 13:896–906, 2020.

[2] R. T. Eastman, J. S. Roth, K. R. Brimacombe, A. Simenov, M. Shen, S. Patnaik, and M. D. Hall. Remdesivir: A review of its discovery and development leading to emergency use authorization for treatment of COVID-19. *ACS Cent Sci*, 6:672–683, 2020.

[3] Y. Xu, O. Barauskas, C. Kim, D. Babusis, E. Murakami, D. Kornyeyev, G. Lee, G. Stepan, M. Perron, R. Bannister, B. E. Schultz, R. Sakowicz, D. Porter, T. Cihlar, and J. Y. Feng. Off-target *in vitro* profiling demonstrates that remdesivir is a highly selective antiviral agent. *Antimicrob Agents Chemother*, 65:e02237–20, 2021.

[4] C. N. Arachchige, L. A. Prendergast, and R. G. Staudte. Robust analogs to the coefficient of variation. *J Appl Stat*, 0:1–23, 2020.
