## Supplementary Materials Figure 1 for "Remdesivir to treat COVID-19: can dosing be optimized?"

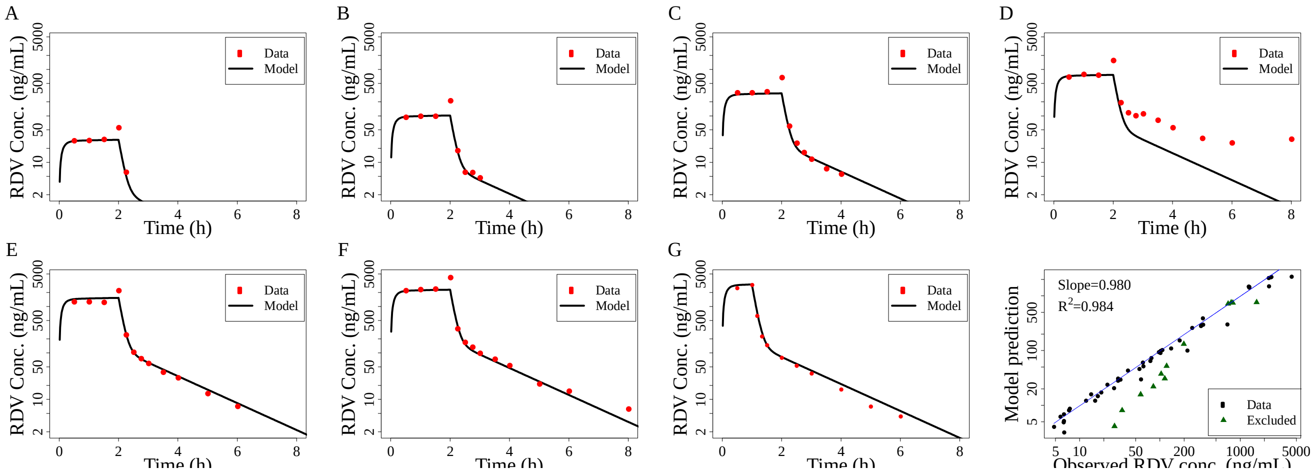


**Figure S1.** Median RDV plasma concentration from single dose experiments (Humeniuk et al. 2020, main text ref. 15), with different RDV dose sizes and infusion durations, compared to our pharmacokinetic model prediction (Equation (1), main text). (**a**) 3 mg dose for a 2 h infusion; (**b**) 10 mg dose for a 2 h infusion; (**c**) 30 mg dose for a 2 h infusion; (**d**) 75 mg dose for a 2 h infusion; (**e**) 150 mg dose for a 2 h infusion; (**f**) 225 mg dose for a 2 h infusion; (**g**) 150 mg dose for a 1 h infusion (first administration of RDV from a multi-dose study). (**f**) Linear regression comparing the model predictions with the data to demonstrate how well the model explains the data. Data from 75 mg dosing excluded in the fitting are marked in green.
