## Supplementary Materials Figure 2 for "Remdesivir to treat COVID-19: can dosing be optimized?"

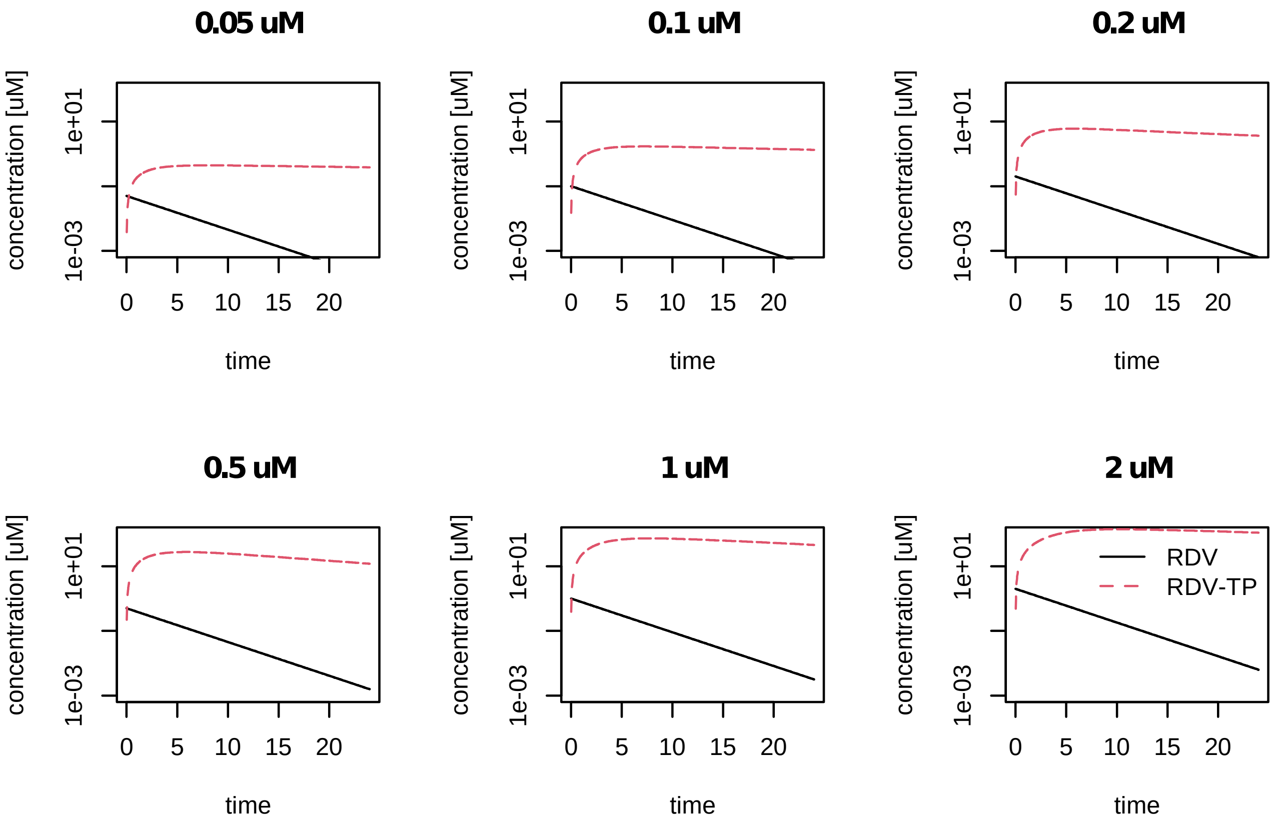


**Figure S2**. Extracellular RDV concentrations and intracellular RDV-TP concentrations during in vitro dose-response experiments. This figure shows the concentrations of RDV and RDV-TP when an initial RDV concentration is supplied in cell culture medium. The parameters are the same as in Figure 2C and D. The title indicates the initial RDV concentration (in µM), the black lines the RDV concentration over time, the red lines the intracellular RDV-TP concentrations over time. We assume that the RDV concentration exponentially declines at 37 °C as described in Ayateneo et al. 2020 (main text ref. 38).
