## Supplementary Materials Table 1 for "Remdesivir to treat COVID-19: can dosing be optimized?"

**Table S1.** Pharmacokinetic parameters for metabolite RDV-TP from single-dose experiments from Humeniuk et al. 2020 (main text ref. 15), with different RDV dose size and infusion durations, compared to our pharmacokinetic model prediction (equation (1), main text). Pharmacokinetic parameters are given in terms of mean and %CV, with the exception of the half-life $t_{1/2}$ for which uncertainty is explained via IQR. Model predictions extending beyond the %CV/IQR are italicized.

| **Dosing** | **Pharmacokinetic parameter (units)** | **Median (%CV or IQR)** | **Model prediction** |
|---|---|---|---|
| 75 mg infused over 2 hours | $C_{max}$ (μM) | 2.5 (16.2) | 2.5 |
| | $C_{24}$ (μM) | 2.2 (23.3) | 1.67 |
| | $AUC_{\infty}$ (h·μM) | 176 (31.1) | 184 |
| | $t_{1/2}$ (h) | 42.7 (30.6-47.4) | *48.0* |
| 150 mg infused over 30 minutes | $C_{max}$ (μM) | 6.0 (46.1) | 8.79 |
| | $C_{24}$ (μM) | 3.7 (40.9) | 4.0 |
| | $AUC_{\infty}$ (h·μM) | 297 (28.3) | 381 |
| | $t_{1/2}$ (h) | 36.0 (27.3-41.5) | 34.4 |
| 75 mg infused over 2 hours | $C_{max}$ (μM) | 5.9 (37.7) | 5.5 |
| | $C_{24}$ (μM) | 3.3 (55.7) | 2.0 |
| | $AUC_{\infty}$ (h·μM) | 394 (49.9) | 291 |
| | $t_{1/2}$ (h) | 49.0 (26.6-69.5) | 39.4 |
